# The Mediating Effect of Social Support on the Relationship Between Cognitive Frailty and Depression in Older Adults

**DOI:** 10.1101/2025.10.19.25338323

**Authors:** Hao Guo, Zhuoyue Xue, Linping Shang, Yuling Li

## Abstract

**Objective:** To examine the mediating effect of social support between Cognitive Frailty and depression in older adults, thereby providing evidence for targeted interventions to mitigate both conditions.

**Methods:** Using data from the 2018 Chinese Longitudinal Healthy Longevity Survey (CLHLS), a sample of 4,589 non-demented older adults aged 65 years and above was selected to investigate the relationships among social support, cognitive frailty, and depression. A simple mediation analysis model was employed to analyze the mediating effect of social support on the relationship between cognitive frailty and depression.

**Results:** The prevalence of Cognitive Frailty was 27.82%, and the detection rate of depressive symptoms was 15.14%. Cognitive frailty was positively correlated with depression (*r*=0.164, *P*<0.001) and negatively correlated with social support (*r*=−0.036, *P*<0.05). Social support was also negatively correlated with depression (*r*=−0.100, *P*<0.001). The direct and total effects of cognitive frailty on depression were statistically significant (95% CI: [1.276, 1.824] and [1.308, 1.858]), and the mediating effect of social support between cognitive frailty and depression was statistically significant (95% CI: [0.006, 0.063]).

**Conclusion:** Cognitive frailty can exacerbate depressive symptoms in older adults, but this relationship can be partially mitigated by enhancing social support. Comprehensive measures should be implemented to improve the level of social support for older adults, which may help delay the progression of cognitive decline and reduce the severity of depression.

## 1. Introduction

With China’s rapid economic and social development, the process of population aging is accelerating. Depression is a common psychological issue prevalent among middle-aged and older adults. Depression may lead to conditions such as cardiovascular disease^[1]^, immune dysfunction^[2]^, and cognitive decline^[3]^. Cognitive frailty is defined as the co-occurrence of physical frailty and cognitive impairment. Diagnosing cognitive frailty requires the exclusion of concurrent Alzheimer’s disease or other forms of dementia, as well as the determination of the severity and type of cognitive impairment. Cognitive frailty is a risk factor for disability, poor quality of life, dementia, and mortality^[4]^, and is also a significant risk factor for depression^[5]^. Depression and cognitive frailty significantly impair older adults’ quality of life, imposing substantial healthcare and socioeconomic burdens. Research indicates a negative correlation between social support and depression^[6]^, highlighting its positive effects on both physical and mental health^[7]^. Social support also serves as a key predictor of cognitive frailty in older adults^[5]^. Given the strong interrelationships among these three factors, this study aims to clarify whether social support mediates the relationship between cognitive frailty and depression in Chinese older adults, and to explore the structural relationships among them. The findings will guide the implementation of effective clinical interventions, thus providing a reference for the prevention and improvement of depression and the delay of cognitive frailty in older adults.

## 2. Data and Methods

### 2.1 Data Source

The Chinese Longitudinal Healthy Longevity Survey (CLHLS) is a national longitudinal study that tracks the health status, longevity, and influencing factors among older adults. Participants primarily comprise individuals aged 65 and above, covering both urban and rural areas nationwide. This study utilized the 2018 survey from this database as its research sample, initially including 15,410 non-demented individuals aged 65 years or older. After filtering for missing and anomalous values in relevant variables, a final sample of 4,589 elderly individuals was ultimately included. See Figure 1 for details. The latest available data in the database is the 2018 survey.

**Figure 1.**
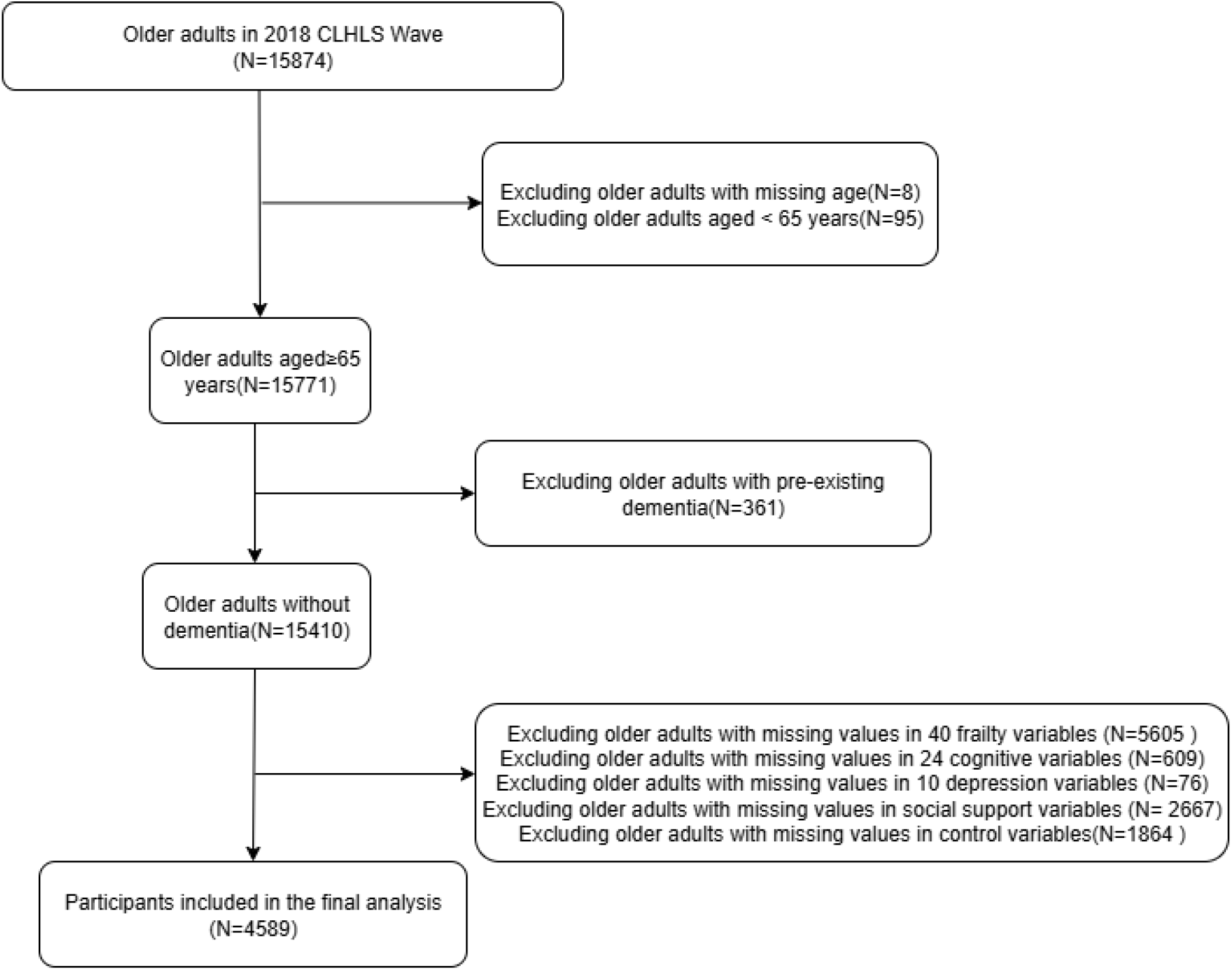
Screening process

### 2.2 Ethical Considerations

The CLHLS has received approval from the Ethics Committee of Peking University (IRB No.: IRB00001052-13074), and all original participants provided written informed consent. This was a retrospective study based on survey data from the CLHLS. The data were accessed for research purposes starting from October 2024. The authors did not have access to information that could identify individual participants during or after data collection as the dataset was fully de-identified.

### 2.3 Measures

#### 2.3.1 Control variables

Building upon prior literature^[8, 9]^, the relevant covariates included in this study comprised demographic characteristics (age, gender, current residence, living alone status, educational attainment, pre-retirement occupation, income level) and lifestyle factors (smoking, alcohol consumption, physical activity). “Living with family” and “residing in a care home” were categorized as non-living alone. Educational attainment was measured by “total years of schooling completed,” with a threshold of one year used for categorization. Income status was assessed using the question “How would you describe your standard of living compared to others in your area?” and categorized into three levels: affluent, ordinary, and impoverished.

#### 2.3.2 Explanatory Variables

Cognitive Frailty was defined as the co-occurrence of physical frailty and cognitive impairment, after excluding dementia. In this study, cognitive frailty in older adults was constructed using the following CLHLS measures. First, dementia was excluded using the question “Do you have dementia?” Cognitive function was assessed using the MMSE scale, which demonstrated an internal consistency coefficient (Cronbach’s α) of 0.893. The scale comprises 24 items, with a maximum total score of 30 points. This study employed reverse scoring: the original MMSE score was subtracted from the total possible score of 30 points. Scores of 0–3 indicated good cognitive function, while scores of 4-30 indicated cognitive impairment. Physical frailty^[8, 10-12]^ was primarily assessed using the Frailty Index (FI), which comprises 40 questions covering basic activities of daily living (ADL), instrumental activities of daily living (IADL), functional limitations, self-rated health status, visual and hearing impairment, chronic disease status, major illnesses in the past two years, and investigator-rated health status. Each negative response was scored as 1 point, and each positive response as 0 points. The FI was calculated by summing individual scores and dividing by the total possible score. Following classification methods in other literature^[13]^, the frailty index was categorized as follows: FI ≤ 0.21 indicates no frailty, and FI > 0.21 defines a frail state.

#### 2.3.3 Mediating Variable

Social support is a beneficial, intrinsic characteristic of social relationships that provides individuals with emotional, instrumental, or informational support. Based on a literature review^[8, 14, 15]^, social support was constructed through three dimensions: family support (financial assistance from children, caregivers during physical discomfort, seeking help when facing difficulties, regular conversation partners, and confidants), community support (social services provided by the community), and governmental support (pension insurance and medical insurance). The total social support score ranged from 0 to 19.

#### 2.3.4 Dependent Variable

The CLHLS used the CESD-10 scale to assess depression levels among older adults. This scale showed high reliability, with a Cronbach’s alpha coefficient of 0.927. The depression scale comprises ten questions. The six response options (“always,” “often,” “sometimes,” “rarely,” “never,” and “cannot answer”) for each question were assigned a score from 0 to 3 points^[16]^ Questions 5, 7, and 10, which pertain to positive psychological states, employed reverse scoring, while the remaining seven questions used forward scoring. The scores from all ten questions were summed, yielding a maximum total of 30 points. A score ≥ 10 was defined as indicative of depressive symptoms^[17]^.

### 2.4 Statistical Methods

Statistical analysis was conducted using SPSS version 25.0. Demographic and health data related to depressive status were analyzed using independent samples *t*-tests for continuous measures and chi-square tests for categorical measures. Continuous variables are presented as mean ± standard deviation (SD), while categorical variables are expressed as frequencies and proportions. Spearman correlation analysis was employed to examine the relationships among cognitive frailty, depression, and social support in older adults, specifically determining the strength and direction of linear associations between variables. The mediation analysis was conducted using the SPSS PROCESS macro (Model 4) to test whether the relationship between cognitive frailty and depression was mediated by social support. The mediation model was established if the 95% confidence interval (CI) for the indirect effect (a×b) did not include zero. Bootstrap testing was used to validate the mediating role of social support between cognitive frailty and depression in older adults. *P*< 0.05 was considered statistically significant.

## 3. Results

### 3.1 General Characteristics of Participants

A total of 4,589 participants were included in the analysis, consisting of 1,941 males (42.3%) and 2,648 females (57.7%). The mean age was (84.76 ± 11.92) years. Residence distribution: urban 2,298 (50.1%), rural 2,291 (49.9%); Living alone: 780 (17.0%) yes, 3,809 (83.0%) no; Educational attainment: <1 year 2,419 (52.7%), ≥1 year 2,170 (47.3%). Pre-retirement occupation: Professional work 263 (5.7%), Non-professional work 4,326 (94.3%). Income status: Affluent 770 (16.8%), Ordinary 3,295 (71.8%), Poor 524 (11.4%). Smoking status: Yes 745 (16.2%), No 3,844 (83.8%). Alcohol consumption: Yes 693 (15.1%), No 3,896 (84.9%). Exercise habits: Yes 1,342 (29.2%), No 3,247 (70.8%). Cognitive frailty: Yes 1,277 (27.8%), No 3,312 (72.2%). Mean Social support score was (8.03 ± 2.65), with sub-scores: Family support (5.08 ± 1.37), Community support (1.68 ± 2.08), and Government support (0.89 ± 0.31).

### 3.2 Comparison of Depressive Symptom Prevalence by Characteristics

As shown in Table 1, 695 participants (15.14%) exhibited depressive symptoms, while 3,849 (84.86%) did not. Among those with depressive symptoms, participants were significantly more likely to be female, have lower educational attainment, engage in non-professional occupations, possess a relatively poor economic status, engage in less physical exercise, and exhibit cognitive frailty (*P*< 0.001). Compared to non-depressed elderly individuals, those with depressive symptoms exhibited significantly lower levels of social support (*P*< 0.001).

**Table 1.**
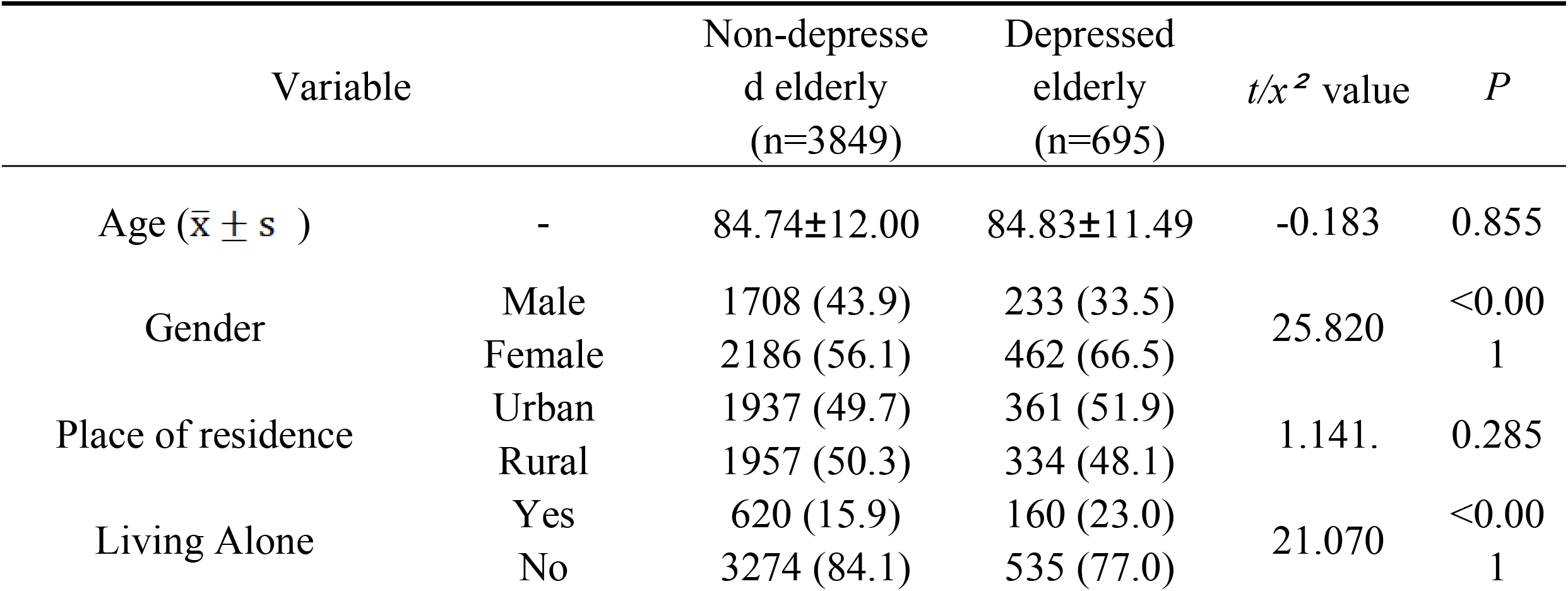
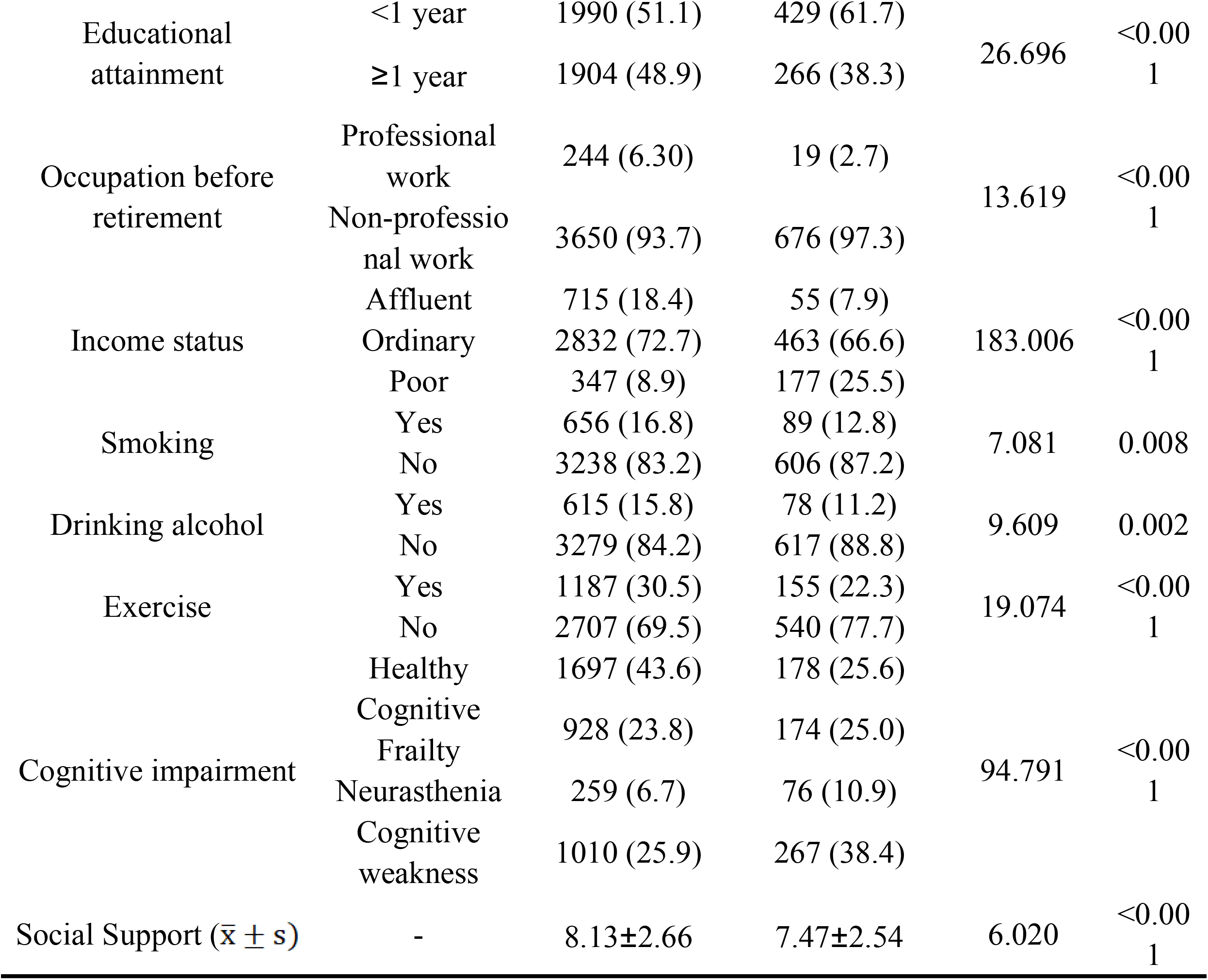
Basic characteristics and differential analysis of depressive symptoms in the elderly [n(%)]

### 3.3 Correlation Analysis of Cognitive Frailty, Social Support, and Depression in Older Adults

After controlling for demographic variables, Spearman correlation analysis revealed that cognitive frailty was positively correlated with depression (*r*=0.164, *P*<0.001). Social support exhibited negative correlations with both cognitive frailty (*r*=-0.036, *P*<0.05) and depression (*r*=-0.100, *P*< 0.001). See Table 2.

**Table 2.**
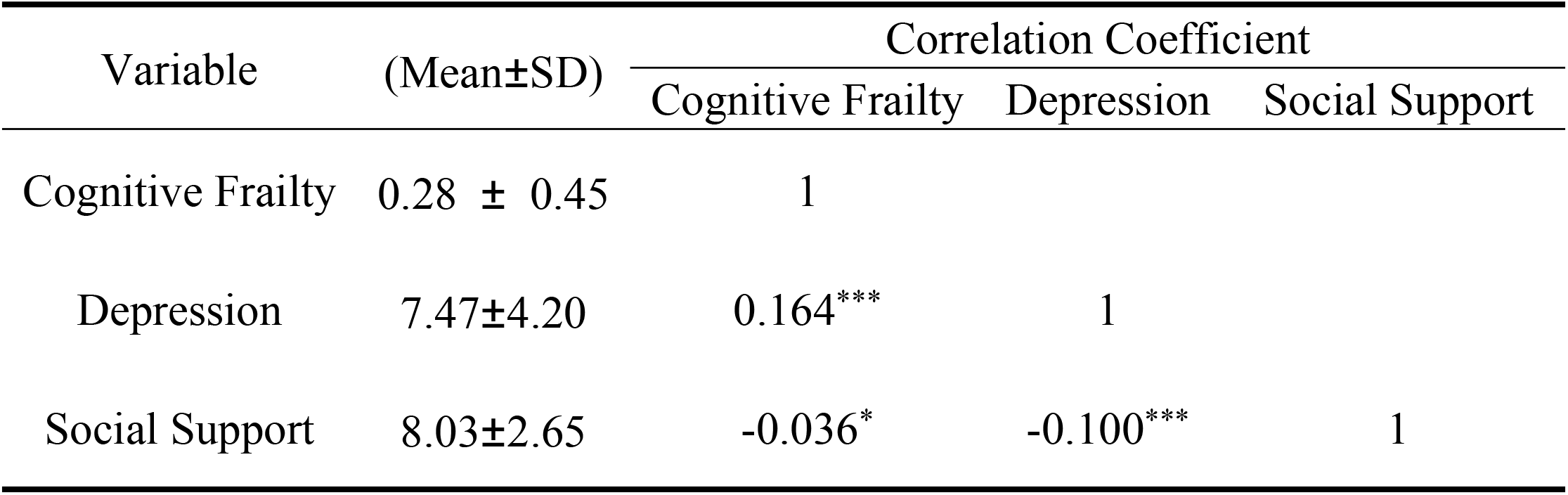
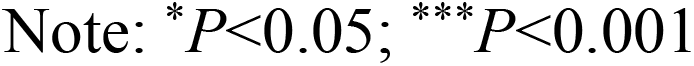
Correlation analysis of cognitive frailty, social support, and depression in older adults.

| Variable | (Mean±SD) | Correlation Coefficient |  |  |
| --- | --- | --- | --- | --- |
|  |  | Cognitive Frailty | Depression | Social Support |
| Cognitive Frailty | 0.28 ± 0.45 | 1 |  |  |
| Depression | 7.47±4.20 | 0.164*** | 1 |  |
| Social Support | 8.03±2.65 | -0.036* | -0.100*** | 1 |

Note: \* $P < 0.05$ ; \*\*\* $P < 0.001$

### 3.4 Testing the Mediating Effect of Social Support between Cognitive Frailty and Depression

After controlling for relevant covariates, mediation analysis was conducted using the PROCESS macro (Model 4). Regression results (Table 3) indicate that cognitive frailty exerts a significant negative influence on social support (*t* = -2.437, *P*< 0.05) and a significant positive influence on depression (*t* = 11.092, *P*< 0.001). Social support exerts a significant negative effect on depression (*t* = -6.461, *P*< 0.001). The mediating effect of social support between cognitive frailty and depression was further examined using the bias-corrected percentile bootstrap method. As shown in Table 4, the direct and total effects of cognitive frailty on depression were statistically significant (95% CI: 1.276–1.824; 95% CI: 1.308–1.858), and the mediating effect of social support between cognitive frailty and depression was also statistically significant (95% CI: 0.006–0.063). The direct effect value was 1.550, the total effect value was 1.583, and the mediating effect value was 0.032. All three effects were significant, and the indirect effect (a×b) and direct effect (c’) shared the same sign. Therefore, social support partially mediated the relationship between cognitive frailty and depression. See Figure 2.

**Table 3.** Regression analysis results for variable relationships in the mediation effect model.

| Regression equation |  | Overall fit indices |  |  | Regression Coefficient Significance |  |
| --- | --- | --- | --- | --- | --- | --- |
| Outcome variable | Predictor Variable | $R$ | $R^2$ | $F$ | $\beta$ | $t$ |
| Social Support | Cognitive Frailty | 0.143 | 0.021 | 10.683 | -0.228 | -2.437* |
| Depression | Cognitive Frailty | 0.362 | 0.131 | 69.023 | 1.550 | 11.092*** |
|  | Social Support |  |  |  | -0.142 | -6.461*** |

Note: \* $P < 0.05$ ; \*\*\* $P < 0.001$ , all tests two-tailed

**Table 4.**
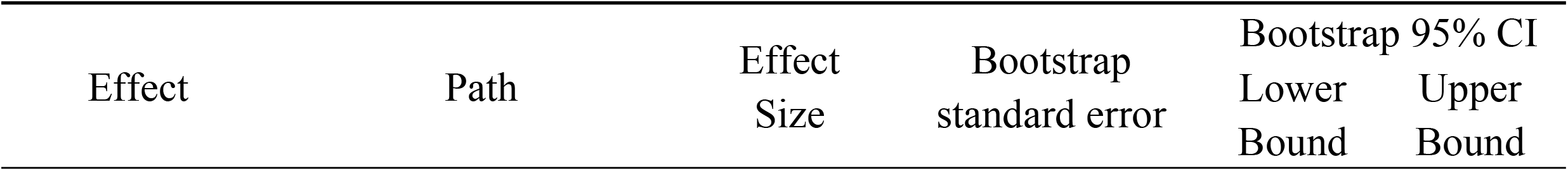
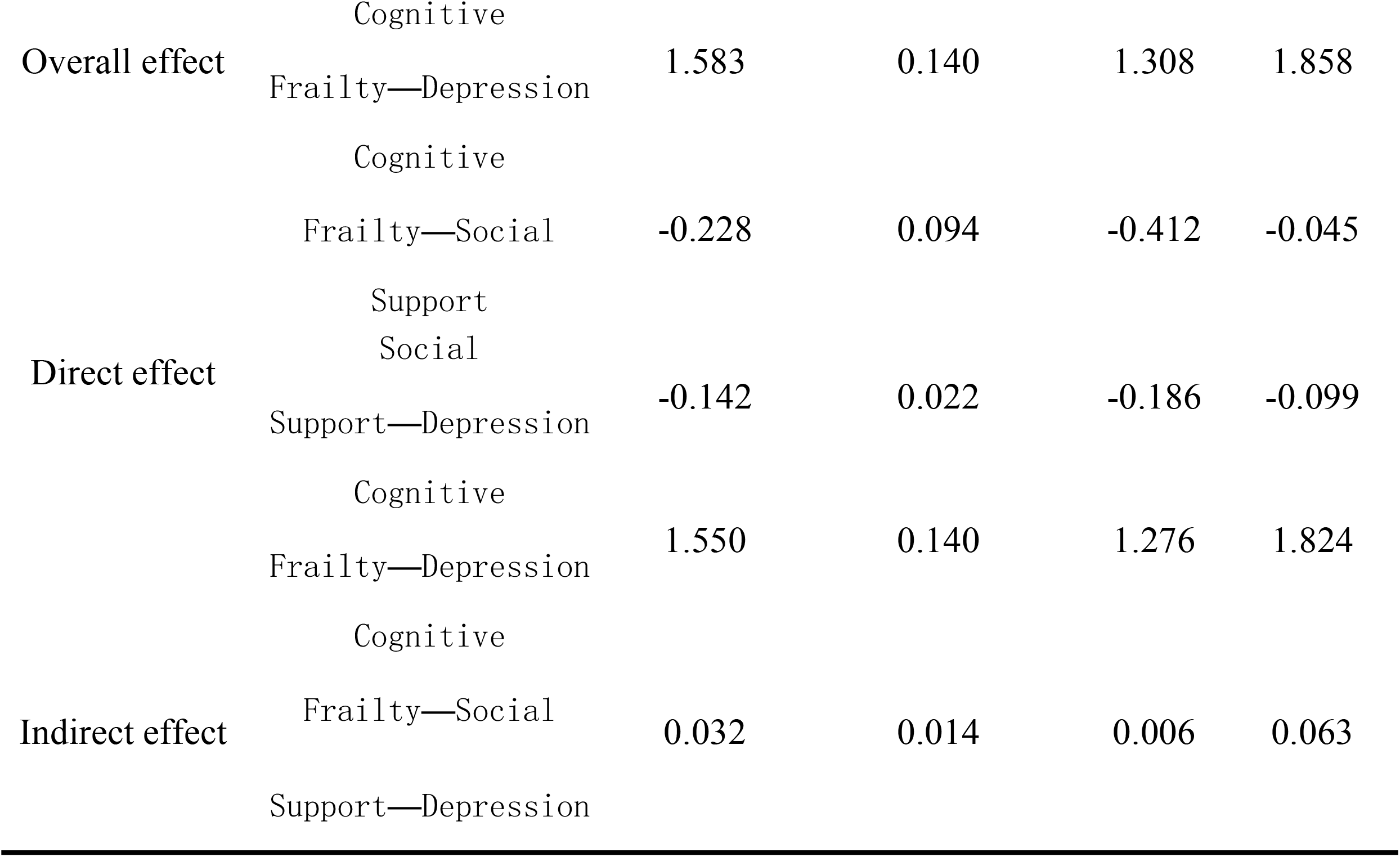
Bootstrap mediation effect test.

**Figure 2.**
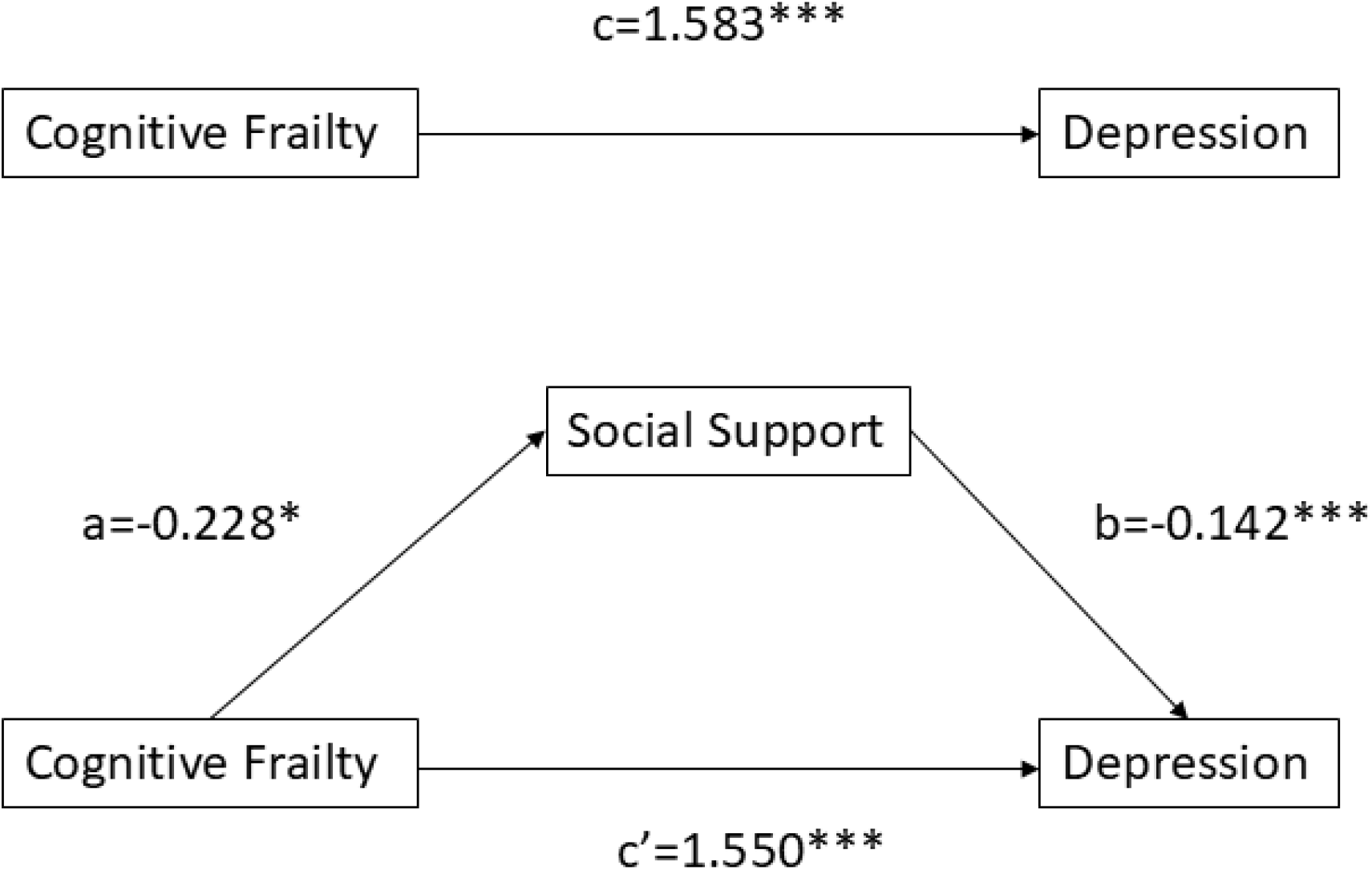
Mediating model and regression coefficient of social support in relation to cognitive frailty and depressive symptoms Note: \**P*<0.05; \*\*\**P*<0.001

## 4. Discussion

### 4.1 Higher Prevalence of Cognitive Frailty Among Depressed Older Adults

In this study, the detection rate of depression was 15.14%, comparable to previously reported prevalence rates among Chinese older adults, 12.84%^[18]^. Regarding cognitive frailty, the detection rate was 27.8%, consistent with findings from Zheng Jing et al. (26.2%^[19]^). This study further revealed that 61.6% of depressed elderly individuals exhibited cognitive frailty, representing a relatively high prevalence. A meta-analysis^[20]^ indicates that the overall prevalence of depression among individuals with cognitive frailty is 46%. Cognitive frailty and depression in older adults are closely interrelated. Previous studies^[21-24]^ have demonstrated that both frail older adults and those with cognitive frailty face an elevated risk of developing depression. Furthermore, compared to those without depressive symptoms, individuals with depression exhibit an increased vulnerability to frailty ^[25]^. To further validate the relationship between the two, Deng et al.^[26]^ employed a bidirectional Mendelian Randomization (MR) study to observe that a bidirectional causal relationship does indeed exist between frailty and depression. Simultaneously, depressive symptoms are associated with cognitive impairment^[3]^, which can also lead to the onset of frailty. The combination of frailty and cognitive impairment results in the development of cognitive frailty.

### 4.2 Correlation Analysis of Cognitive Frailty, Social Support, and Depression

Research indicates that social support exhibits a negative correlation with both cognitive frailty and depression, consistent with prior findings^[5, 6]^. This suggests that adequate social support may help mitigate the severity of cognitive frailty and depressive symptoms in older adults, thereby underscoring the importance of social support in preventing and alleviating both conditions. Social support provides cognitive stimulation through activities such as interaction with others, sharing experiences, and acquiring new knowledge. Such stimulation helps maintain brain activity and preserve normal cognitive function. Prolonged stress, anxiety, and depression can impair the immune system, leading to reduced immune function. Conversely, social support offers emotional reassurance and encourages healthy lifestyle choices among older adults, thereby contributing to physical well-being and maintaining normal immune system function, which in turn may slow the progression of cognitive frailty. Furthermore, social support encourages older adults to participate in social activities, expand their social circles, and strengthen their social support networks. This reduces feelings of loneliness and social isolation, thereby alleviating depressive symptoms. Research has also identified a significant positive correlation between cognitive frailty and depression. The relationship between cognitive frailty and depression may involve a complex bidirectional process^[26]^. Older adults may experience feelings of helplessness and diminished self-worth due to declining physical function, reduced Activities of Daily Living (ADLs), weakened social functioning, and cognitive decline. This can subsequently lead to psychological issues such as anxiety and depression. Conversely, depression may lead to poor nutritional status, sleep disturbances, and emotional dysregulation, which can severely compromise physical health and contribute to frailty and cognitive impairment. Furthermore, frailty, cognitive function, and depression share overlapping pathophysiological domains and exhibit common susceptibility to relevant stressors, suggesting a potential correlation between cognitive frailty and depression^[8, 27]^.

### 4.3 Social Support Partially Mediates the Relationship Between Cognitive Frailty and Depression

Previous literature extensively demonstrated the correlation between cognitive frailty and depression in older adults^[20, 21, 23]^. However, few studies have explored the role of other variables in this relationship. The findings of this study indicate that social support partially mediates the association between cognitive frailty and depression in older adults. That is, addressing cognitive frailty directly can alleviate depressive symptoms in older adults, or the impact of cognitive frailty on depressive symptoms can be mitigated by enhancing social support levels among this population. First, we must understand the mechanism through which social support operates between cognitive frailty and depression in older adults. As an external resource, social support fulfills individual needs through emotional, informational, and instrumental assistance, thereby reducing negative emotions and strengthening psychological resilience. This, in turn, influences cognitive frailty and depressive mood. Among the elderly, as cognitive function gradually declines and frailty deepens, depressive mood becomes more prevalent. Social support may serve a connecting and buffering role in this context. Findings indicate that social support may partially mediate the relationship between cognitive frailty and depression through pathways such as alleviating loneliness, providing emotional support, and cognitive stimulation. Furthermore, analysis of mediating effects must acknowledge that mediation does not represent a simple causal relationship but involves complex interactions among multiple variables. This study, which included only social support as a variable, should also consider other potential influencing factors such as an individual’s psychological state, health status, and quality of daily living. Furthermore, as this study employs a cross-sectional design, consideration should be given to conducting longer-term longitudinal research to further validate the mediating effect and sustained influence of social support between cognitive frailty and depression in older adults. Interdisciplinary collaboration is encouraged, bringing together experts and practitioners from the fields of social support, cognitive frailty, and depression research to collectively explore how to maximize the role of social support in managing the mental health of older adults.

In response to these findings, it is recommended that government departments formulate and implement policies to encourage and support social organizations and volunteer groups in providing social support services for older adults. Increased funding should be allocated to establish and refine a comprehensive social support service system for older adults, alongside monitoring mechanisms to ensure service quality and effectiveness. Community organizations may conduct social support activities for older adults, such as regular gatherings and mental health lectures, providing emotional and informational support. Concurrently, family members and the wider community should attend to older adults’ mental health needs, offering greater care and support to foster a warm and harmonious social environment. Healthcare institutions may integrate mental health resources to provide counseling and cognitive training services for older adults, while recognizing the role of social support in mental well-being. It is recommended that healthcare institutions and community service providers regularly assess older adults’ levels of social support, promptly identify individuals with insufficient support networks, and provide targeted assistance. Educational institutions may develop mental health education programs for older adults, enhancing their awareness of mental health issues and social support mechanisms while fostering positive attitudes towards social support. These efforts will contribute to improving older adults’ mental well-being, enhancing their quality of life and sense of happiness, and actively advancing the development of a harmonious and healthy aging society.

## Data Availability

Tsuyoshi Hamano Kyoto Sangyo Daigaku Humayun Kabir McMaster University Department of Health Research Methods Evidence and Impact Fabíola Bof de Andrade Fundacao Oswaldo Cruz Instituto Rene Rachou Ioannis Liampas Panepistemio Thessalias Tmema Iatrikes.

https://opendata.pku.edu.cn/dataverse/CHADS

